# Predicted long-term impact of COVID-19 pandemic-related care delays on cancer incidence and mortality in Canada

**DOI:** 10.1101/2021.08.26.21261149

**Authors:** Talía Malagón, Jean H.E. Yong, Parker Tope, Wilson H. Miller, Eduardo L. Franco, for the McGill Task Force on the Impact of COVID-19 on Cancer Control and Care

## Abstract

**Objectives:** The COVID-19 pandemic has affected cancer care worldwide. This study aimed to estimate the long-term impacts of the pandemic on cancer incidence and mortality in Canada using a mathematical model.

**Methods:** We developed a stochastic microsimulation model to estimate the cancer care disruptions and its long-term impact on cancer incidence and mortality in Canada. The model reproduces cancer incidence, survival, and epidemiology in Canada, by using cancer incidence, stage at diagnosis and survival data from the Canadian Cancer Registries. We modeled reported declines in cancer diagnoses and treatments recorded in provincial administrative datasets from March 2020-June 2021. We assumed that diagnostic and treatment delays lead to an increased rate of death. Based on the literature, we assumed each 4-week delay in diagnosis and treatment would lead to a 6% to 50% higher rate of cancer death. Results are the median predictions of 10 stochastic simulations.

**Findings:** The model predicts that cancer care disruptions during the COVID-19 pandemic could lead to 21,247 (2·0%) more cancer deaths in Canada in 2020-2030, assuming treatment capacity is recovered to 2019 pre-pandemic levels in 2021. This represents 355,172 life years lost expected due to pandemic-related diagnostic and treatment delays. The highest absolute expected excess cancer mortality was predicted in breast, lung, and colorectal cancers, and in the provinces of Ontario, Québec, and British Columbia. Diagnostic and treatment capacity in 2021 onwards highly influenced the number of predicted cancer deaths over the next decade.

**Interpretation:** Cancer care disruptions during the Covid-19 pandemic could lead to significant life loss; however, most of these could be mitigated by increasing diagnostic and treatment capacity in the post-pandemic era to address the service backlog.

**Funding:** Canadian Institutes of Health Research

**Research in context:** *Evidence before this study:* We performed a review of modeling studies predicting the impact of pandemic-induced disruptions to cancer care on cancer survival outcomes. We searched MEDLINE on 2 July 2021 for records published from 1 January 2020 with no language restrictions. Our search consisted of index keywords [*Cancer* AND *COVID-19* AND [(*delay* AND *diagnosis*) OR (*delay* AND *screening*) OR (*delay* AND *treatment*)] AND *outcomes* AND *modelling study*]. We identified 14 studies that model the long-term effect of disruptions to cancer screening programs, diagnostic intervals, and to treatment intervals for common cancers. Most studies (9/14) assessed the impact of cancer screening disruptions but did not assume any treatment disruptions. Disruptions to cancer screening services in high income health systems were estimated to lead to small increases in cancer incidence and mortality, even with immediate resumption of screening to services after disruption periods. Fewer studies examined the impact of diagnostic referral and treatment disruptions; these are similarly predicted to lead to increases in cancer incidence and mortality, with varying impacts depending on cancer site. Due to difficulties in obtaining real-time healthcare data, previous studies have relied on assumptions regarding the duration of health care disruptions (1-, 3-, 6-, 12-, to 24-months) rather than on empirical data. All studies restricted their analysis to the impact on a single or a few cancer sites.

*Added value of this study:* Our stochastic microsimulation model is the first to assess the population-level impact of diagnostic and treatment disruptions on overall cancer mortality across all sites. Using Canadian cancer statistics and expert validation of treatment modalities, we constructed a model that reproduced pre-pandemic cancer mortality data. An important added value of this analysis compared to previous studies was that we were able to integrate empirical data on cancer-related procedures during the pandemic era to model disruptions to cancer care.

*Implications of all the available evidence:* We estimate there could be a 2·0% increase over expected cancer mortality between 2020-2030 in Canada due to pandemic-related disruptions to diagnostic and treatment intervals. Our results identified that a 10-20% increase in cancer care service capacity over pre-pandemic levels could prevent a considerable amount of the predicted excess cancer-related deaths by reducing diagnostic and treatment backlogs. By stratifying our reported outcomes by sex, age, province, and cancer site, we provide a long-term perspective that can inform post-pandemic public health policy or aid in prioritization of patients in the event of a resurgence of COVID-19. While our model is specific to Canada, it could be applied to countries that have experienced comparable COVID-19-related healthcare disruptions.

## Introduction

The COVID-19 pandemic has led to severe disruptions in health care delivery worldwide. Cancer patients are at particularly high risk of negative outcomes from delays in diagnosis and treatment. A meta-analysis estimated that each 4-week delay in cancer surgery increases the rate of mortality by 6-8% across several major cancer sites.^1^ The pandemic has impacted the entire cancer care trajectory due to changes in healthcare seeking behavior and health system capacity.^2^ Many individuals have delayed consulting for cancer-related symptoms due to access barriers, or fears of infection with SARS-CoV-2 in a healthcare setting.^3,4^ Healthcare staffing shortages may occur due to redeployment to the pandemic response, leave due to SARS-CoV-2 infection, burnout, or increased childcare responsibilities. Key cancer diagnostic tests such as endoscopies, colonoscopies, mammographies, computerized tomography (CT) scans, and magnetic resonance imaging (MRI) scans have declined,^5,6^ either due to fewer patient referrals or delays in accessing these services. Many cancer surgeries and other treatments were postponed or delayed during the pandemic. ^2^

In Canada, both public health and cancer services are publicly funded and provincially managed. In March 2020, most provinces declared a state of public health emergency, which led to cancellation and postponement of many cancer treatments, screenings, and routine health care visits. Provincial health agencies eventually issued directives and practice guidelines for cancer care provision during the pandemic.^7^ Many included criteria for prioritization and triage of cancer patients in case of healthcare disruptions, and specified cancer treatments should be maintained as a high priority service. Cancer treatments gradually resumed over the next few months,^8^ and our discussions with officials from cancer agencies indicated that most perceived cancer service provision had returned to normal over the course of 2020. However, fewer cancer treatments were performed in 2020,^8^ suggesting many cancer cases experienced delays in their diagnosis and treatment due to the pandemic.

Our objective was to predict the long-term impact of pandemic-related cancer diagnostic and treatment delays on cancer mortality in Canada by cancer site, age, sex, and province. We also examined factors which could mitigate or increase the expected cancer mortality over the next decade.

## Methods

We built an individual-level stochastic microsimulation model in C++. The model simulates the health trajectories of cancer cases over time and was designed to capture the impacts of diagnostic and treatment delays on cancer incidence and mortality. The model’s main features are described below; a more detailed description of parameters and assumptions can be found in an online supplementary technical appendix at https://doi.org/10.5683/SP2/REMSZ6.

### Model structure

We modeled the incidence of 25 cancer sites (bladder, brain, breast, central nervous system, cervix, colorectal, esophagus, Hodgkin lymphoma, kidney & renal pelvis, larynx, leukemia, liver, lung, melanoma, multiple myeloma, non-Hodgkin lymphoma, oral, ovary, pancreas, prostate, stomach, testes, thyroid, uterus, and all other cancers) based on 2015-2017 Canadian incidence rates^9^ and population size by sex and age.^10^ Cancers were assigned an overall TNM stage at diagnosis based on the distribution reported by provincial cancer registries.^11^

Two death dates were sampled for each cancer case: their expected cancer death date and their expected death date from other causes. The expected cancer death date was sampled from a net cancer survival function depending on sex, age, cancer site, and stage.^12–14^ The expected death date from other causes was sampled from 2017-2019 Canadian life tables by sex,^15^ adjusted for seasonality using a sinusoidal function fitted to weekly death counts. A cancer case’s actual death date was the earliest of these two expected death dates. If the expected cancer death date was earlier than the expected death date from other causes, then we assumed that the person died from cancer.

Four cancer treatment modalities were included in the model: surgery, radiotherapy, chemotherapy, and other. Patients could receive combinations of surgery, radiotherapy, and chemotherapy. Patients in the other category were those who did not receive any of these three treatments and received other care. We were unable to identify a nationally representative dataset for the distribution of cancer treatments in Canada. We based treatment distributions on expert opinion from an Expert Advisory Group of 31 oncologists and surgeons, supplemented with data on treatment distributions from England and the USA.^16,17^ The experts used a survey tool to validate whether the treatment distributions from England and the USA were applicable to Canada given their experience. If they judged treatment probabilities to differ, they were asked to quantify the probability of receiving each treatment for a given cancer site by stage. We combined the probability distributions elicited from experts using an equal weights linear opinion pool.^18^

We defined the treatment interval as the time between the diagnosis date and the start date of curative or palliative treatment. The treatment interval was explicitly modeled: upon diagnosis, each cancer case was assigned a treatment interval sampled from a Weibull distribution, fitted to pre-pandemic reported times to cancer treatment.^6,19^ We defined the diagnostic interval as the time between the date a patient first notices cancer symptoms and the date of diagnosis. This definition included the patient interval^20^ (time between noticing symptoms and presenting to healthcare) in the diagnostic interval, because declines in cancer diagnoses were due to patients both taking longer to present to healthcare as well as a reduction in diagnostic activities during the pandemic. This diagnostic interval was not explicitly modeled; however, the date of diagnosis could be delayed as described below, implicitly increasing the diagnostic interval.

### Pandemic-related diagnostic & treatment delays

We modeled the pandemic’s impact on health care delivery as a relative change in the monthly number of diagnoses and treatments compared to the numbers expected had there been no pandemic. This relative change set the maximum diagnostic and treatment capacity each month. Diagnoses and cancers treatments scheduled to occur which exceeded the monthly system capacity were placed on a backlog. During each time step, the model would attempt to clear the backlog by randomly selecting from the list of undiagnosed cases which were diagnosed, and selecting from the list of diagnosed untreated cases which received treatment, up to the maximum capacity. The backlogs increased during time steps where diagnostic and treatment volumes were below expected levels. Because selection of cases on the backlog was random, some cancer cases did not experience any delays, while other cases could experience significant diagnostic and treatment delays.

Relative changes in surgeries were based on the volume of surgeries per month in 2020-2021 relative to the same month in 2019. Data for surgeries was extracted for all provinces, except Québec, from the Canadian Institute of Health Information (CIHI) web portal up to February 2021, and supplementary data was provided for Québec by the Ministère de la santé et des services sociaux (MSSS) up to March 2021. The estimated relative changes in cancer surgeries are shown in Figure 1A. Declines in cancer surgeries roughly coincided with peaks in COVID-19 hospitalizations in Canada (Figure 1B).^21^ Most provinces experienced a third infection wave around April 2021; we therefore assumed that the trends observed in January-February 2021 would be repeated in March-April 2021.

**Figure 1.**
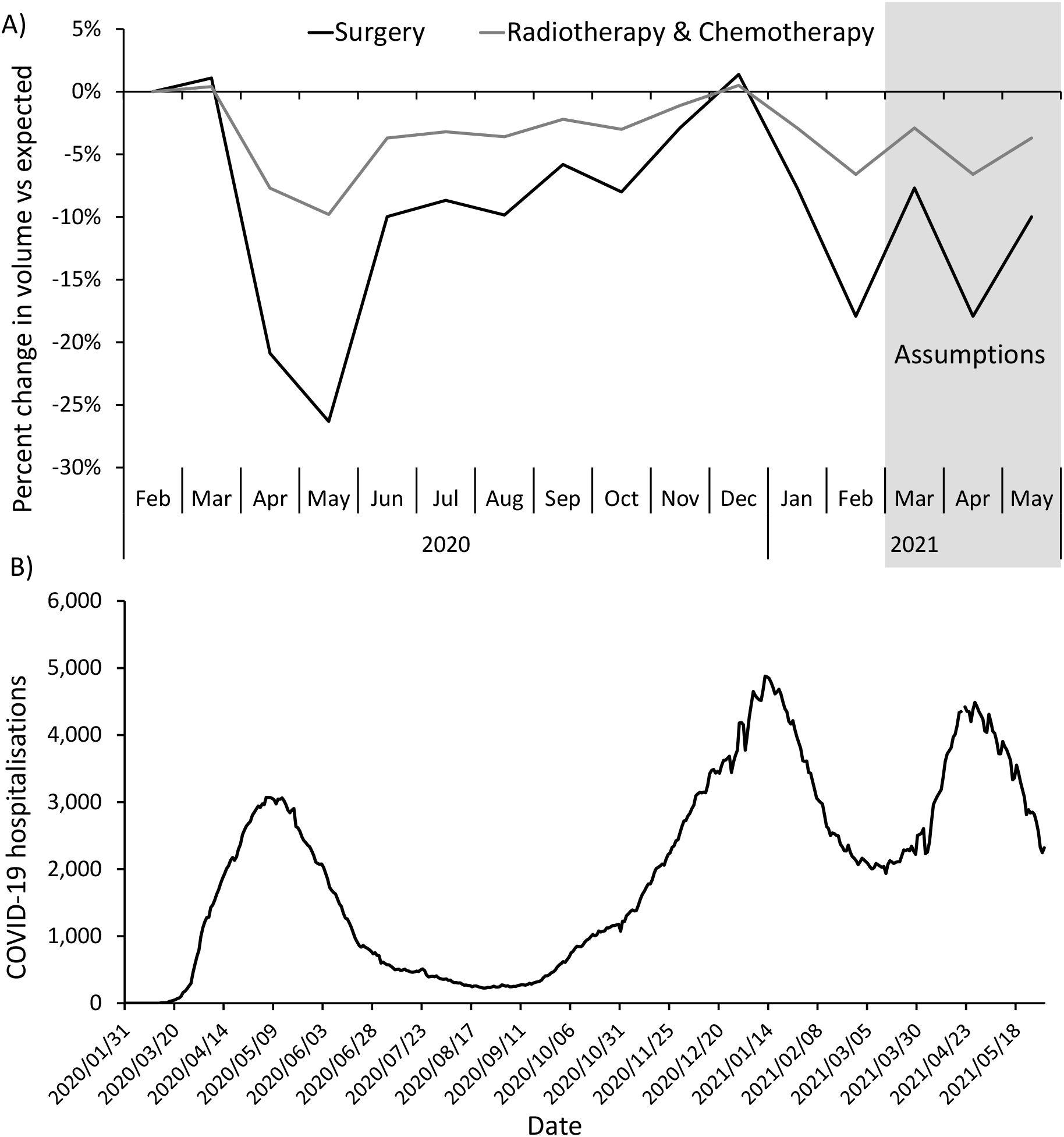
Impact of the COVID-19 pandemic on cancer treatments and hospitals in Canada. A) Modeled percent change in cancer treatments for Canada as a whole. The percent change in surgeries is based on data on the volume of cancer surgeries in 2020-2021 relative to the same month in 2019, using data from the Canadian Institute of Health Information portal extracted on May 28^th^ 2021. The percent change in radiotherapies is based on the yearly volume of radiotherapies in 2020 relative to 2019 reported by the Canadian Institute of Health Information,^6^ rescaled by month using the surgery data. Chemotherapies were assumed to follow the same changes as radiotherapies. B) Number of people hospitalized for COVID-19 in Canada from February 2020 to May 2021. Data compiled by Radio-Canada extracted on June 1^st^ 2021.^21^ AB=Alberta; BC=British Columbia; MB=Manitoba; NB=New Brunswick; NL=Newfoundland & Labrador; NS=Nova Scotia; ON=Ontario; PEI=Prince Edward Island; QC=Québec; SK=Saskatchewan.

There was a small −2.5% yearly percent change in the number of radiotherapies in Canada in 2020 compared to 2019 reported to CIHI.^6^ We rescaled this percent change per month in the model so that the largest declines occurred in Spring, with recovery in the Summer and Fall. Changes in radiotherapies were only modeled for Québec, Ontario, New Brunswick, and Prince Edward Island, as other provinces did not report declines in radiotherapies.^6^ There was no data for chemotherapies; we assumed these would experience the same monthly percent changes as radiotherapies.

We used 2019-2021 monthly pathology report data provided by the MSSS as a proxy for new diagnoses in Québec. This province reported a similar overall percent change in pathology reports as in cancer surgeries over 2020-2021. Based on this, we assumed for other provinces that the monthly decline in new diagnoses would be the same as the reported monthly decline in cancer surgeries (Figure 1A).

The diagnostic delay was the difference between the date the cancer would have been diagnosed without the pandemic, and the actual diagnosis date because of the diagnostic backlog. The treatment delay was the difference between the initially scheduled treatment date, and the actual treatment date because of the treatment backlog. The sum of the diagnostic and treatment delays constituted the total delay experienced by a cancer case. The total delay impacted a cancer’s survival probability by increasing the rate of cancer mortality. A meta-analysis estimated that each 4-week delay in cancer surgery leads to a 1·06-1·08 hazard ratio increase in mortality across several major cancer sites.^1^ Hazard ratios for radiotherapy and systemic therapy delays were more variable, but included similar values in their confidence intervals. Based on this, we assumed that all cancer sites and all treatment modalities would have a 1·06 times higher cancer mortality hazard rate per 4-week total delay. The relationship between time and mortality rates was assumed to be log-linear, with each 4-week delay increasing the mortality rate multiplicatively. The effects were also multiplicative across treatments. Consequently, patients with longer delays and who require multiple treatments were those most likely to experience higher mortality.

For each patient experiencing pandemic-related delays, the model resampled a new cancer death date for them based on their cancer survival function given their total delay. Cancer cases whose resampled cancer death dates were earlier than their original expected death dates were reassigned the earlier death date; the rest retain their original expected death date. This reflects an assumption that delays are either detrimental or have no effect on cancer survival.

### Scenarios and statistical analyses

A counterfactual no-pandemic scenario was used as a comparator. In the base case pandemic scenario, we assumed there would be declines in diagnoses and treatments from March 2020 to May 2021 (Figure 1A), and that, starting June 2021, treatment capacity would return to normal pre-pandemic levels and diagnostic capacity would increase by 15% due to increased diagnostic activities and increased patient interactions with the healthcare system. In sensitivity analyses, we evaluated more pessimistic scenarios, where treatment capacity remains 10-20% below pre-pandemic levels throughout 2021, and more optimistic scenarios, where treatment capacity is increased over normal capacity after June 2021 to decrease the diagnostic and treatment backlogs. We also varied the mortality hazard ratio associated with a 4-week delay due to the high uncertainty regarding the effect of delays on cancer mortality.

Results are the median (minimum-maximum interval) of 10 stochastic simulations per scenario. Excess cancer deaths were calculated as the difference in the number of cancer deaths between pandemic scenarios and the no pandemic counterfactual scenario. Life years lost were calculated as the difference between a case’s initially sampled death date and their resampled death date after delays.

### Validation

The model reproduces net cancer survival by cancer site in Canada and the pre-pandemic (2017-2019) number of cancer deaths reported per year by province (technical appendix Figures 6-8).

## Results

Figure 2 shows the median predicted monthly number of cancer incidence and deaths in Canada for all cancer sites combined between 2017-2030. Cancer incidence and deaths are predicted to increase over time even in a no-pandemic counterfactual due to population ageing and increasing population size. In all pandemic scenarios, a temporary decline in cancer incidence is expected during 2020-2021 due to pandemic-related declines in new diagnoses. Most missed diagnoses are expected to be caught up later, however, as diagnostic capacity recovers and patients return to normal healthcare seeking behaviors. The model predicted an increase in cancer deaths in pandemic scenarios starting from 2020 due to pandemic-related diagnostic and treatment delays.

**Figure 2.**
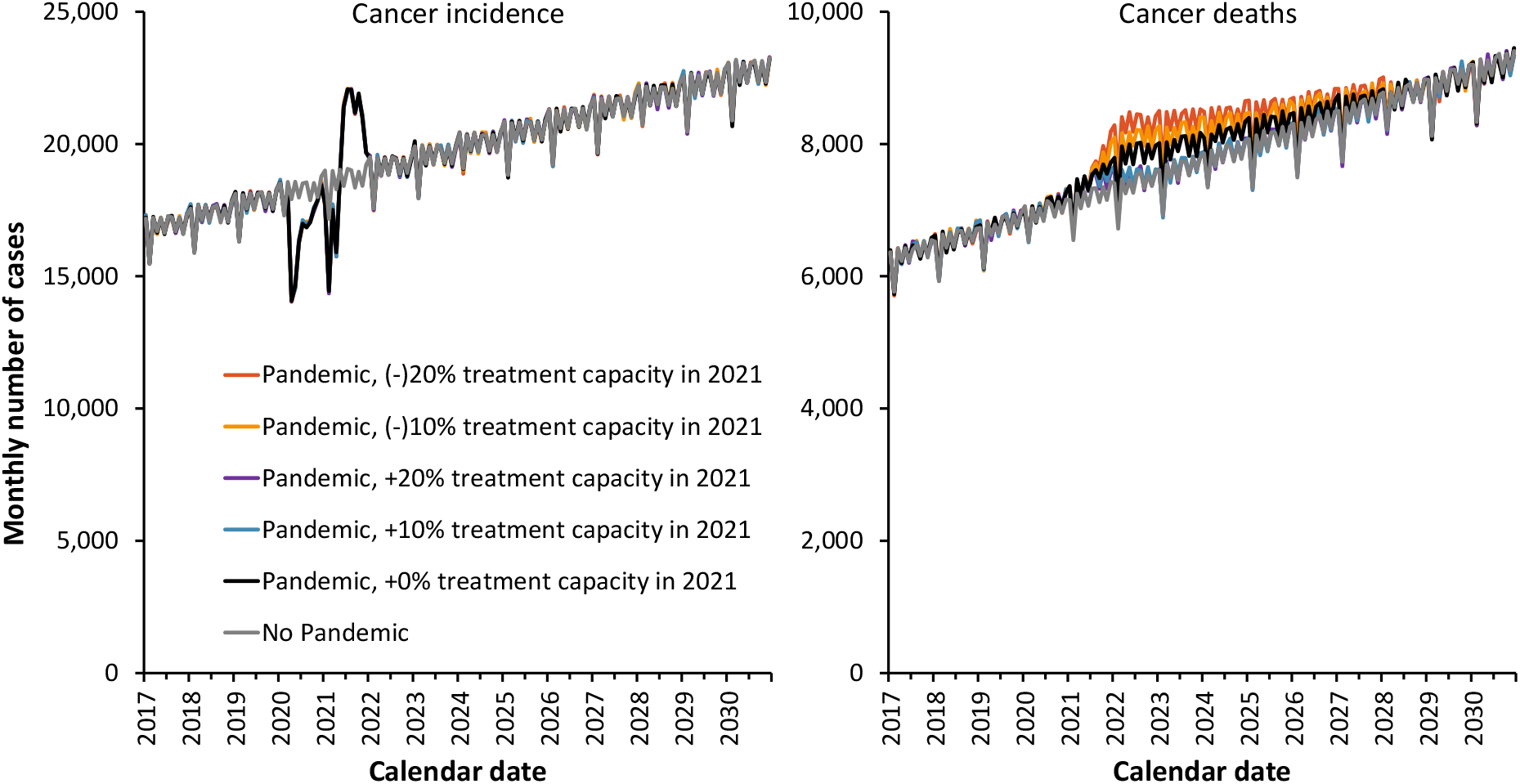
Predicted monthly cancer incidence and deaths for all cancer sites combined, Canada. Results are the median of 10 stochastic simulations for each scenario. The base case scenario is shown in black. All scenarios assume that treatment capacity changes occur starting in June 2021, and that each 4-week delay in cancer care increases the rate of cancer mortality by 6% (hazard ratio of 1.06).

In terms of excess cancer mortality, 21,247 (18,108-26,136) cumulative excess cancer deaths were predicted between 2020-2030 for Canada as a whole in the base case scenario due to pandemic-related delays (Figure 3). This constitutes a 2·0% (1·7%-2·5%) increase over expected cancer mortality over this time period, and 355,172 (348,434-401,887) life years lost. The year with the most excess cancer mortality was predicted to be 2022, with cancer mortality being 6% higher than expected that year. Excess cancer mortality from pandemic-related delays was predicted to last up to 2027, after which the yearly number of cancer deaths would return to expected levels.

**Figure 3.**
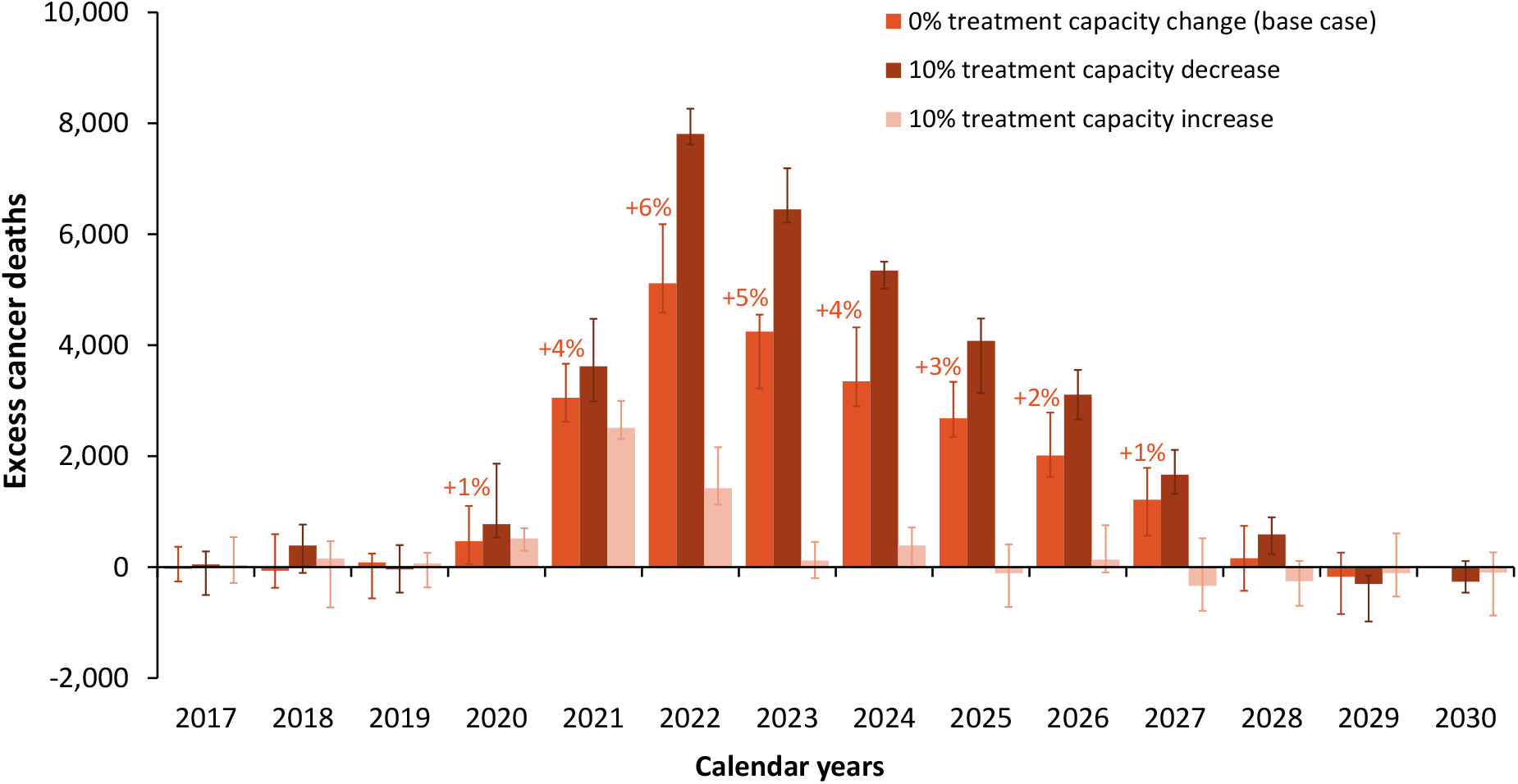
Predicted yearly excess cancer deaths compared those expected without the pandemic for all cancer sites combined, Canada. Results are the median and error bars are the minimum and maximum of 10 stochastic simulations for the base case scenario (recovery in June 2021), and scenarios with ±10% treatment capacity over pre-pandemic levels starting June 2021. Percentages indicate the yearly median relative increase in cancer deaths over expected.

The above results from the base case scenario assume that pre-pandemic cancer treatment capacity levels are recovered by June 2021. However, predicted excess cancer mortality was sensitive to assumed treatment capacity levels in 2021 and after (Figure 2, Figure 3). Continued pandemic-related reductions in treatment capacity would substantially increase excess cancer mortality, while increasing treatment capacity would substantially decrease excess cancer mortality. If capacity for all treatments remains 10% lower than normal throughout 2021, then 33,262 (31,381-35,077) cumulative excess cancer deaths were predicted between 2020-2030. If treatment capacity were increased by 10% over normal, only 4,210 (2,719-5,675) excess cancer deaths were predicted between 2020-2030; this is equivalent to preventing 80% of excess cancer mortality predicted in the base case. Future treatment capacity strongly influenced results because more cancers are expected to be diagnosed and require treatment in 2021/2022 than in 2020 due to diagnostic delays.

Cumulative excess cancer deaths between 2020-2030 predicted in the base case scenario by sex, age, province, and cancer site are presented in Table 1. The relative mortality increase was predicted to be highest in cases diagnosed under age 45, but the highest absolute life years lost were predicted to occur in cases diagnosed between ages 55-74. Most excess cancer deaths were predicted to occur in the most populous provinces of Ontario, Québec, Alberta, and British Columbia. In general, cancer sites with poor expected survival (pancreas, esophagus, lung) were predicted to experience lower relative mortality increases, though lung cancer accounted for many absolute excess deaths due to its high incidence.

**Table 1.**
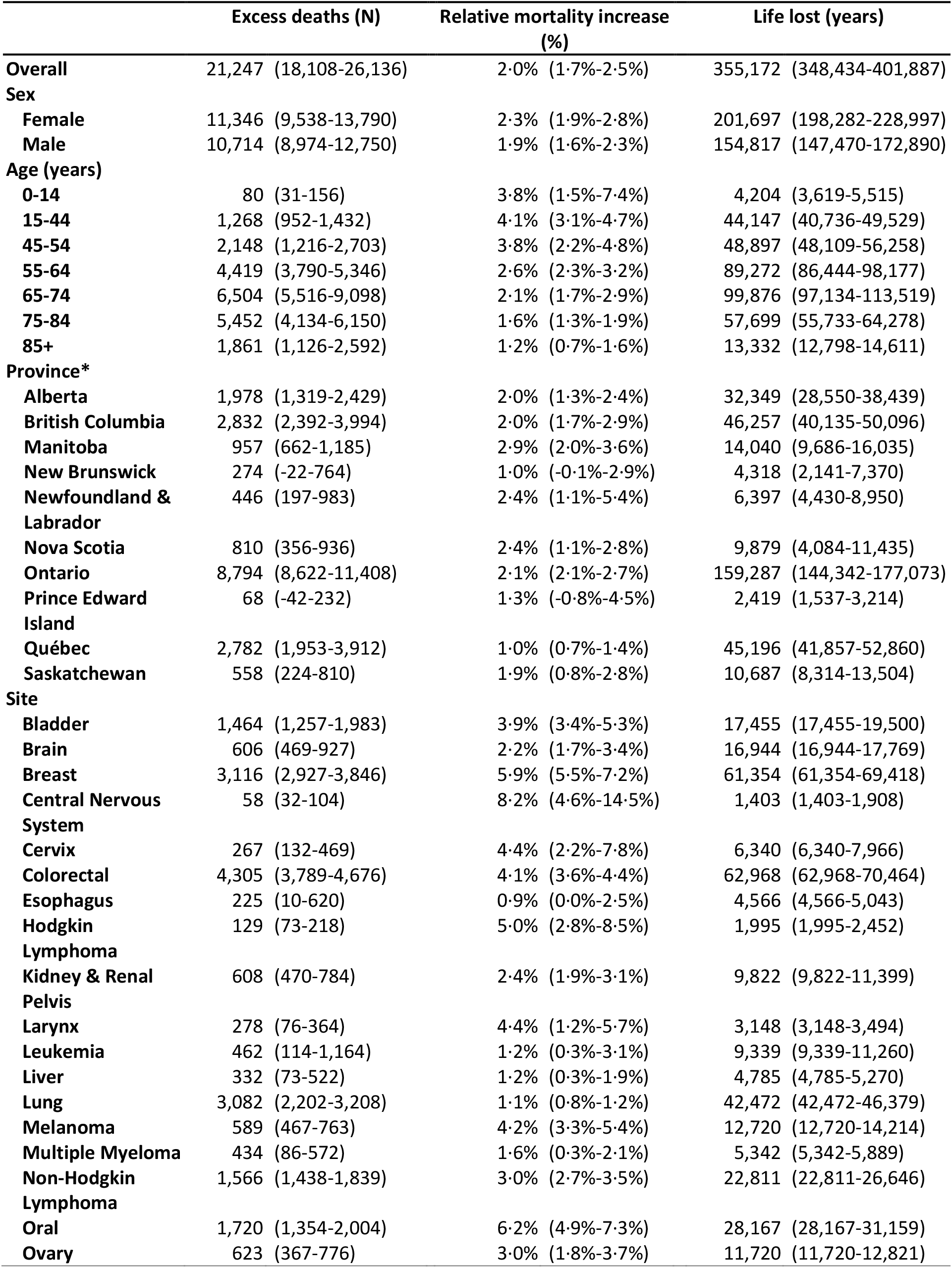
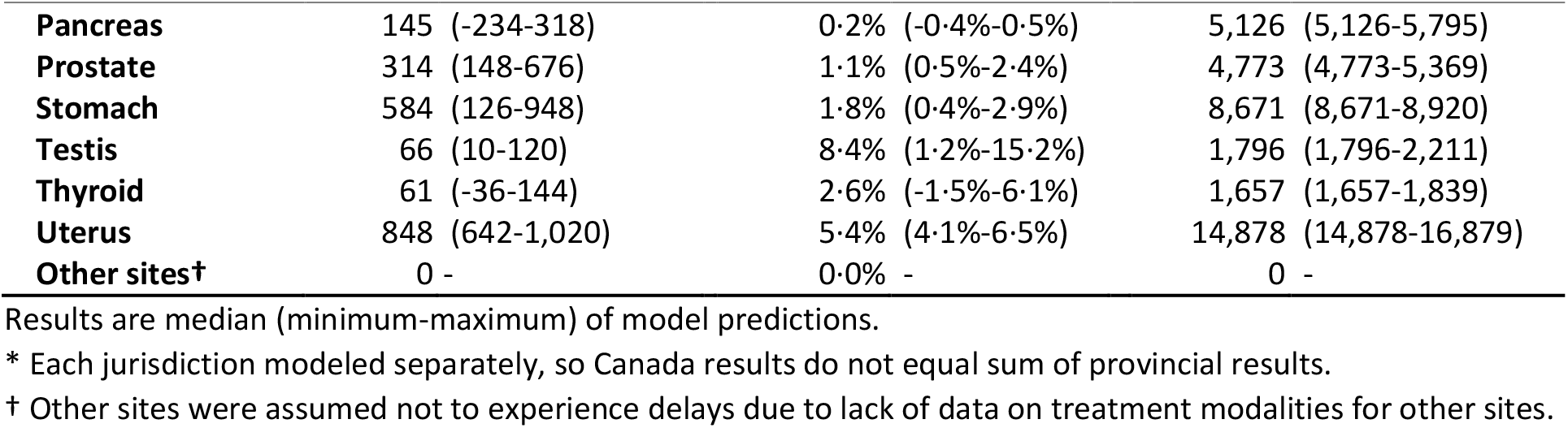
Predicted cumulative excess cancer mortality and life years lost by sex, age, province, and site for Canada 2020-2030 in the base case scenario.

In sensitivity analyses, we varied the effect of delays on the mortality hazard ratio, stratified by site (Table 2). Unsurprisingly, higher hazard ratios led to higher expected excess mortality than in the base case scenario. The time frame for this excess mortality remained the same as in the base case (2020-2027). Further results stratified by province are also be presented through an interactive web application at https://tmalagon.shinyapps.io/CancerCOVID19Model/.

**Table 2.** Predicted cumulative excess cancer mortality by site, assuming different hazard ratios (HR) for the effect of a 4-week total delay on cancer mortality, for Canada 2020-2030.

|  | Excess deaths (N) |  |  |  |  | Relative mortality increase (%) |  |  |  |  |
| --- | --- | --- | --- | --- | --- | --- | --- | --- | --- | --- |
|  | HR 1·03 | HR 1·06* | HR 1·1 | HR 1·2 | HR 1·5 | HR 1·03 | HR 1·06* | HR 1·1 | HR 1·2 | HR 1·5 |
| **Overall** | 12,606 | 21,247 | 31,178 | 42,987 | 59,577 | 1·2% | 2·0% | 2·9% | 4·0% | 5·6% |
| **By Site** |  |  |  |  |  |  |  |  |  |  |
| **Bladder** | 956 | 1,464 | 2,235 | 3,232 | 4,706 | 2·5% | 3·9% | 6·0% | 8·6% | 12·5% |
| **Brain** | 488 | 606 | 824 | 970 | 1,084 | 1·8% | 2·2% | 3·0% | 3·5% | 4·0% |
| **Breast** | 1,953 | 3,116 | 4,735 | 6,932 | 10,478 | 3·7% | 5·9% | 8·9% | 13·0% | 19·7% |
| **Central Nervous System** | 22 | 58 | 94 | 102 | 176 | 3·1% | 8·2% | 13·2% | 14·2% | 24·6% |
| **Cervix** | 183 | 267 | 307 | 498 | 728 | 3·0% | 4·4% | 5·1% | 8·3% | 12·1% |
| **Colorectal** | 2,400 | 4,305 | 5,896 | 8,624 | 12,143 | 2·3% | 4·1% | 5·6% | 8·2% | 11·5% |
| **Esophagus** | 118 | 225 | 300 | 442 | 522 | 0·5% | 0·9% | 1·2% | 1·8% | 2·1% |
| **Hodgkin Lymphoma** | 82 | 129 | 166 | 260 | 362 | 3·2% | 5·0% | 6·4% | 10·1% | 14·1% |
| **Kidney & Renal Pelvis** | 348 | 608 | 866 | 1,304 | 1,960 | 1·4% | 2·4% | 3·4% | 5·1% | 7·7% |
| **Larynx** | 139 | 278 | 354 | 506 | 632 | 2·2% | 4·4% | 5·6% | 8·0% | 9·9% |
| **Leukemia** | 206 | 462 | 832 | 1,198 | 1,686 | 0·5% | 1·2% | 2·2% | 3·1% | 4·4% |
| **Liver** | 294 | 332 | 604 | 554 | 764 | 1·1% | 1·2% | 2·3% | 2·1% | 2·9% |
| **Lung** | 1,574 | 3,082 | 3,791 | 5,116 | 6,405 | 0·6% | 1·1% | 1·4% | 1·9% | 2·3% |
| **Melanoma** | 304 | 589 | 944 | 1,428 | 2,110 | 2·2% | 4·2% | 6·7% | 10·1% | 14·9% |
| **Multiple Myeloma** | 344 | 434 | 560 | 756 | 1,026 | 1·3% | 1·6% | 2·1% | 2·8% | 3·8% |
| **Non-Hodgkin Lymphoma** | 927 | 1,566 | 2,159 | 3,216 | 4,406 | 1·8% | 3·0% | 4·1% | 6·1% | 8·4% |
| **Oral** | 1,164 | 1,720 | 2,440 | 3,159 | 4,087 | 4·2% | 6·2% | 8·9% | 11·5% | 14·8% |
| **Ovary** | 332 | 623 | 882 | 1,226 | 1,326 | 1·6% | 3·0% | 4·3% | 5·9% | 6·4% |
| **Pancreas** | 70 | 145 | 304 | 346 | 560 | 0·1% | 0·2% | 0·5% | 0·5% | 0·9% |
| **Prostate** | 356 | 314 | 502 | 702 | 1,076 | 1·3% | 1·1% | 1·8% | 2·5% | 3·9% |
| **Stomach** | 353 | 584 | 667 | 923 | 1,242 | 1·1% | 1·8% | 2·0% | 2·8% | 3·8% |
| **Testis** | 28 | 66 | 89 | 142 | 177 | 3·6% | 8·4% | 11·3% | 17·9% | 22·4% |
| **Thyroid** | 40 | 61 | 132 | 196 | 256 | 1·7% | 2·6% | 5·6% | 8·4% | 10·9% |
| **Uterus** | 528 | 848 | 1,108 | 1,768 | 2,619 | 3·4% | 5·4% | 7·1% | 11·3% | 16·7% |

HR=hazard ratio. Results are median (minimum-maximum) of model predictions.
\*Base case scenario

## Discussion

In this modeling study, we predicted that diagnostic and treatment delays in cancer care caused by the COVID-19 pandemic would lead to clinically important increases in cancer mortality in Canada. Increasing cancer diagnostic and treatment capacity in the post-pandemic era by ≥ 10% over normal levels was predicted to mitigate a substantial amount of this excess mortality. This is because increased treatment capacity would allow clearing any accumulated treatment backlogs, but also would allow to deal with an expected increased future demand in cancer treatments due to delayed cancer diagnoses during the pandemic. Interestingly, provinces that reported higher declines in cancer treatments were not necessarily those predicted to experience the highest relative increases in cancer mortality. This suggests that age structure and cancer case mix also influence the expected population-level excess cancer mortality. Cancer sites with moderate to high net survival and in younger patients were those predicted to experience the highest relative mortality increases. This may be because cancers with higher survival are those which stand to lose the most from delays; many cancers with low net survival such as pancreatic and esophageal cancers would not be expected to survive very long even with normal wait-times.

Much evidence suggests an important part of the observed decline in cancer treatments is likely attributable to cancer diagnostic delays, and not necessarily treatment delays. Cancer surgical wait-times returned to normal and even improved in Canada during 2020, however the cumulative number of surgeries was lower than in previous years.^5,6^ Fewer pathology reports, MRI scans, and CT scans have been performed than in previous years.^5,6^ Many diagnostic procedures and cancer screenings such as colonoscopies and mammographies were interrupted or slowed due to the pandemic.^5,22^ This suggests there is a backlog of undiagnosed cancers that have yet to be identified for treatment, either due to decreased patient interactions with the health care system or due to delays in the diagnostic pathway. An important part of the model-predicted excess mortality is attributable to these delayed diagnoses which will eventually require treatment. These delayed diagnoses may not yet be apparent in observed treatment backlogs and waitlists, but should be accounted for in the planning for future healthcare needs, especially as many are likely to present with more advanced cancers.

Some provincial governments have announced investments to address diagnostic and treatment backlogs across the healthcare system, but it is unclear whether these will be sufficient to prevent excess cancer mortality. For example, while the government of Ontario has pledged $300M to clear surgical backlogs, the Financial Accountability Office has estimated it would take $1·3B to clear the surgery and diagnostic procedure backlog if hospitals operate at +11% capacity for surgeries and +18% capacity for diagnostic procedures.^23^ Ongoing healthcare worker staffing shortages in many provinces may also limit increases in diagnostic and treatment capacity. While Canada has achieved high COVID-19 vaccination coverage, future epidemic waves caused by new SARS-CoV-2 variants may also lead to further health care disruptions. For these reasons, we have modeled both pessimistic and optimistic scenarios to assess the full range of potential future outcomes.

The causal effect of cancer care trajectory delays on cancer mortality is the most uncertain parameter in our model. Research on this topic has been challenging due to a wait time paradox, a type of confounding by indication where cancers with worse survival have shorter diagnostic and treatment intervals due to being prioritized in cancer care pathways.^24^ We based the effect of delays in the model on results from a high-quality systematic review that accounted for the wait time paradox.^1^ The effects of delays in our model were similar to those assumed by other models. By our calculation, Hartman *et al*. assumed mortality hazard ratios between 1·03-1·2 per 4-week treatment delay based on data from 5 million patients in the US National Cancer Database;^25^ Sud *et al*. assumed mortality hazard ratios between 1·09-1·17 per 4-week diagnostic delay based on their own literature review.^26^ We believe our base case scenario assuming a hazard ratio of 1·06 for both diagnostic and treatment delays provides conservative predictions, given that studies have found many cancer sites and treatment modalities with larger hazard ratios.^1,25,26^ Because the true causal effect of delays remains highly by cancer site, we provided estimates by cancer site assuming different effects of delays (Table 2).

Predictions of the impact of the COVID-19 pandemic on cancer deaths have varied across models.^25–28^ While it is difficult to directly compare models due to structural differences, we believe assumptions regarding the extent and duration of disruptions to cancer-related healthcare greatly influence differences in predictions. Due to the difficulty of obtaining real-time health care data, previous models have mostly examined the impact of hypothetical 3- to 24-month pandemic disruptions to cancer care. For example, we predicted fewer excess breast cancer deaths than Maringe *et al*., who predicted a 8-10% increase over 5 years in England.^28^ This may be because they modeled a complete stop to screening and routine diagnostic investigations over a 12-month period, based on an assumption the pandemic would disrupt health care over a long time. However, we predicted more excess breast cancer deaths than Alagoz *et al*., who predicted a 0.5% increase between 2020-2030 in the United States.^27^ This may be because they modeled a shorter 6-month disruption to screening and few disruptions to treatments, based on an assumed rapid response of health care providers to reinstitute cancer services. Our predictions are based on empirical data on the volume of cancer-related procedures, which suggest that while cancer care provision did rapidly recover in Canada, there have been long-lasting disruptions to the number of cancer surgeries and diagnoses performed since March 2020. While our model does not include the effects of screening, pandemic-related disruptions to screening are also likely to lead to further excess cancer mortality for breast, colorectal, and cervical cancers.^29,30^

Different countries and jurisdictions have been impacted differently by the pandemic, depending on implemented public health measures and the resilience of their health care systems. In Canada, cancer treatments were generally prioritized over many other types of health care, so the declines in cancer care procedures were not as large as they could have been. However, we believe there are some lessons that are broadly generalizable across settings. While our predicted numbers are specific to Canada, we believe our predictions and those of other models are indicative of the magnitude of the effect that pandemic-related healthcare disruptions could be expected to cause in many countries with similar COVID-19 epidemic profiles. Most importantly, our findings highlight that recovering and even increasing cancer treatment capacity in the post-pandemic would help avoid much of the predicted excess cancer mortality.

## Data Availability

The data collected to inform model parameters is reported in a supplementary online technical appendix available at https://doi.org/10.5683/SP2/REMSZ6. The data for model predictions are publicly available as R objects at the McGill University Dataverse (https://doi.org/10.5683/SP2/AQHVJB). A public web application to further explore results by province will be made available at https://tmalagon.shinyapps.io/CancerCOVID19Model/.

https://doi.org/10.5683/SP2/REMSZ6

https://tmalagon.shinyapps.io/CancerCOVID19Model/

## Author contributions

TM conceived and programmed the model, ran simulations, performed analyses, and wrote the first draft of the manuscript. JY helped parameterize the model and validated model assumptions for analyses. WM provided clinical input for model parameterization and assumptions. PT constructed the expert treatment validation survey, extracted data for model parameters, and performed literature reviews. ELF was the principal investigator for the study, obtained funding and provided team supervision and advice. All authors read, provided feedback, and approved the final protocol and manuscript.

## Acknowledgements

We would like to thank Larry Ellison from Statistics Canada for providing data on net cancer survival in Canada; James Jung from the Canadian Partnership Against Cancer for extracting data from the Canadian Institute for Health Information Portal; Elba Gomez Navas from the Canadian Partnership Against Cancer for developing the companion web application for model results; Annie Bourassa, Christiane Bertrand, and Joëlle Sarra-Bournet from the Québec Ministère de la santé et des services sociaux for providing data on cancer surgeries, radiotherapies, and pathology reports in Québec. Parts of this material are based on data and information compiled and provided by Cancer Care Ontario. However, the analyses, conclusions, opinions and statements expressed herein are those of the author, and not necessarily those of Cancer Care Ontario.

